# Safe and effective pool testing for SARS-CoV-2 detection

**DOI:** 10.1101/2021.04.08.20205781

**Authors:** Marie Wunsch, Dominik Aschemeier, Eva Heger, Denise Ehrentraut, Jan Krüger, Martin Hufbauer, Adnan S. Syed, Gibran Horemheb-Rubio, Felix Dewald, Irina Fish, Maike Schlotz, Henning Gruell, Max Augustin, Clara Lehmann, Rolf Kaiser, Elena Knops, Steffi Silling, Florian Klein

**Author notes:** **Corresponding author:** Florian Klein. These authors contributed equally to this article (Steffi Silling, Florian Klein).

## Abstract

**Background / Objectives:** The global spread of SARS-CoV-2 is a serious public health issue. Large-scale surveillance screenings are crucial but can exceed diagnostic test capacities. We set out to optimize test conditions and implemented high throughput pool testing of respiratory swabs into SARS-CoV-2 diagnostics.

**Study design:** In preparation for pool testing, we determined the optimal pooling strategy and pool size. In addition, we measured the impact of vortexing prior to sample processing, compared pipette- and swab-pooling method as well as the sensitivity of three different PCR assays.

**Results:** Using optimized strategies for pooling, we systematically pooled 55,690 samples in a period of 44 weeks resulting in a reduction of 47,369 PCR reactions. In a low prevalence setting, we defined a preferable pool size of ten in a two-stage hierarchical pool testing strategy. Vortexing of the swabs increased cellular yield by a factor of 2.34, and sampling at or shortly after symptom onset was associated with higher viral loads. By comparing different pooling strategies, pipette-pooling was more efficient compared to swab-pooling.

**Conclusions:** For implementing pooling strategies into high throughput diagnostics, we recommend to apply a pipette-pooling method, using pool sizes of ten samples, performing sensitivity validation of the PCR assays used, and vortexing swabs prior to analyses. Our data shows, that pool testing for SARS-CoV-2 detection is feasible and highly effective in a low prevalence setting.

## 1. Introduction

The SARS-CoV-2 pandemic is a serious public health problem of unprecedented magnitude in recent times. In particular individuals at older ages or with comorbidities are at a high risk to develop an acute respiratory distress syndrome (ARDS) requiring hospitalization and intensive care [1]. Therefore, it is essential to control person-to-person transmission in order to protect vulnerable individuals and limit the number of severe cases. Until herd immunity is achieved by vaccination, nonpharmaceutical interventions need to be applied. Many countries could successfully contain the spread of COVID-19 through social distancing or lock-down measures, contact tracing, quarantine, and large-scale testing in the ongoing pandemic [2,3]. In order to control viral transmission when lifting lock-down strategies, large-scale testing and surveillance are critical interventions. These approaches are based on frequent tests of individuals e.g. by rapid antigen-based tests or reverse transcription-real-time PCR (rRT-PCR) to detect SARS-CoV-2 in swab specimens. However, large-scale surveillance screenings can exceed the test capacities of diagnostic laboratories.

Pooling swab specimens for PCR testing can increase test capacities and limit the consumption of reagents [4]. Pool testing is highly efficient in a setting of low disease prevalence and the availability of highly sensitive test methods [5]. It can be applied to enable surveillance screenings of asymptomatic individuals in public institutions e.g. hospitals, schools or retirement homes, which carry a high risk for superspreading events and severe disease courses. When pool testing is established, test conditions need to be optimized including (a) the type of pooling strategy and pool sizes, (b) sample preparation and pooling method, (c) the quality of SARS-CoV-2 detection by PCR. In this study, our aim was to determine and implement the optimized pool testing procedure into the diagnostic routine for SARS-CoV-2 detection.

## 2. Materials and Methods

### 2.1 Pool testing algorithm

Pooling efficiency was computed using a web tool published by Bilder and colleagues [5,6]. Calculations were performed assuming a test sensitivity of 99% or 95% and a test specificity of 99%. The expected number of tests was computed for different pool sizes as described [5].

### 2.2 Swab specimens

Oropharyngeal or combined nasal/oropharyngeal swabs were collected and transferred into MSwab™ Medium, UTM-RT/mini (COPAN Diagnostics, Murrieta, CA), BD ESwab™ (Becton & Dickinson, Sparks, MD, USA), Sigma Transwab® Purflock® (Medical Wire & Equipment, Corsham, Wiltshire, England), or PBS. All specimens were processed at the Institute of Virology, University Hospital of Cologne within 12 hours after collection. Samples were stored for validation procedures at - 80°C.

### 2.3 Samples, clinical data and Ethics

All samples and clinical data were collected on the wards or outpatient departments of the University Hospital of Cologne. No identifying data were used for the patient’s characterization. According to §15 subparagraph 3 (Professional Code for Physicians in Germany) ethical principles of WMA Declaration of Helsinki were respected.

The study was approved by the Institutional Review Board (Ethics Committee) of the Medical Faculty, University Hospital Cologne, Germany (ethical vote no. 20-1638).

### 2.4 Sample preparation and pooling

After arrival in our laboratory, samples were vortexed 5 seconds, and preselected to be tested individually or in pools. To determine the cellular content of the same specimens before and after vortexing, human β-globin-gene quantification was performed as published [7].

For the pipette-pooling method and to simulate various pool sizes, positive specimens with various Ct-values were combined with increasing volumes of negative samples combined as a stock, accordingly. For the swab-pooling method, nine SARS-CoV-2 negative and one positive swab were used. After removal of the transport medium, 1.2 ml PBS was added to the tube containing the swab and vortexed 5 seconds. The PBS was then transferred to the next swab tube and vortexed. Following this principle, ten swabs were merged. Preparation time was measured.

### 2.5 Reverse transcription, amplification and detection with three instruments

(Instrument I) Nucleic acid extraction of 500 µl sample volume was performed using the MagNA Pure® 96 DNA and Viral NA Large Volume Kit eluted in 100 µl elution buffer, followed by amplification on LightCycler® 480 II (Roche Diagnostics, Mannheim, Germany) according to the manufacturer’s instructions. Detection of SARS-CoV-2 was conducted using the RealStar® SARS-CoV-2 RT-PCR Kit 1.0 (altona Diagnostics, Hamburg, Germany) or LightMix® SarbecoV E-gene plus equine arteritis virus (EAV) control (TibMolBiol, Berlin, Germany). (Instrument II) Processing of swabs was implemented with Panther Fusion® Hologic® and SARS-CoV-2 was detected using 5 µl of total RNA in 20 µl of LightMix® SarbecoV E-gene plus β-globin as internal control (TibMolBiol, Berlin, Germany). As second target the N-gene was amplified (inhouse primer sets in multiplex PCR, data unpublished). (Instrument III) Detection of SARS-CoV-2 was performed on a Roche cobas® 6800 using the cobas® SARS-CoV-2 kit targeting the E-gene/ORF-1a/b regions according to manufacturer’s protocol. SARS-CoV-2 was quantified using dilution series of cell culture supernatant extrapolated to approved standards (INSTAND e.V., Düsseldorf, Germany), measured on all instruments, and Ct-values adjusted to Instrument III (Ct_a_).

### 2.6 Data analysis

For correlation analysis, a spearman’s rank correlation was used. For comparing β-globin-gene concentrations, a Mann-Whitney test was performed. To assess statistical differences in Ct-values comparing pooling methods or PCR assays, a multiple comparison one-way ANOVA was used. For comparing preparation times and for matched-pair analysis, a paired t-test was used. The amplification factor was calculated as published [8], Kruskal-Wallis test was performed. GraphPadPrism 7.0 (GraphPad Software, Inc.) was used for statistical analysis. Figures were created using Adobe Illustrator 18.1 (Adobe Inc.).

## 3. Results

### 3.1 Hierarchical pool testing

Pool testing can be performed using different strategies. In this study we conducted two-stage hierarchical pooling procedures (Figure 1A). Swab samples were combined and tested in a single PCR reaction. If the pool test was positive, the remaining sampling material of the included specimens was retested separately to detect the infected individual. If the pool test was negative, all individuals were declared as not infected [5,9,10].

**Figure 1.**
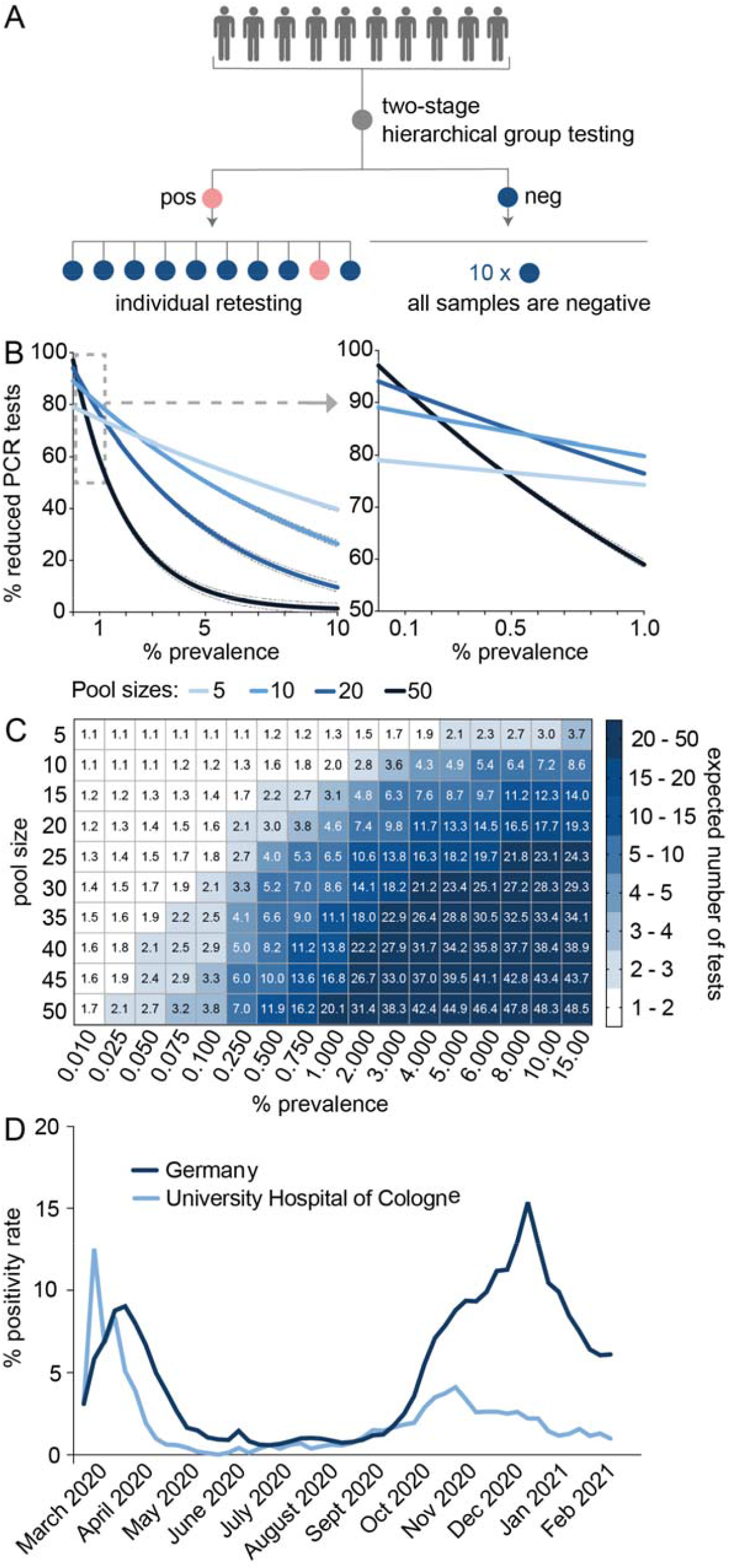
Hierarchical pool testing for SARS-CoV-2 detection. A: Illustration of the two-stage hierarchical pool testing strategy. B: The reduction of PCR tests compared to individual testing (continuous lines are nonlinear regression curves, outer dotted lines are 95% or 99% test sensitivity, respectively) and C: the expected number of tests for different pool sizes are shown. Calculations were performed as published [6,9]. D: The mean positivity rate per week of tests performed at the University Hospital of Cologne and in Germany (as published [11]) are shown.

Pool testing efficiency depends on the disease prevalence. Bilder and colleagues [5,6] proposed an algorithm to compute the expected number of tests when performing two-stage hierarchical pool testing (Figure 1B and C). As the disease prevalence increases, the reduction of PCR tests declines due to the retesting of individual samples of positive pools. However, the pooling efficiency of smaller pool sizes declines more slowly compared to pooling 20 or more samples.

At the time pool testing was initiated, the positivity rate at the University Hospital of Cologne was 3.88% (Figure 1D). However, by excluding samples of symptomatic individuals and recently positive tested persons, the positivity rate of pooled samples was below 0.1%.

### 3.2 Pooling method and sensitivity of the PCR assays

Pool testing requires optimal sample conditions in order to minimize false negative results. Pre-analytic factors can influence the test results. We could not detect a significant difference in viral loads indicated by Ct-values comparing oropharyngeal swabs with combined nasal/oropharyngeal swabs. However, samples taken in the late phase of infection had significantly lower viral loads compared to earlier timepoints (Supplementary Figure 2A and B). Therefore, pool testing needs to be sensitive and safe in detecting even low virus concentrations. We investigated whether vortexing increases the number of cells released from the swabs (Figure 2A). We measured the β-globin-gene concentration in the medium of n=33 swabs without vortexing and of the same specimens after 5 seconds of vortexing. The average increase of β-globin concentration after vortexing was 2.34-fold (95% CI 1.622-3.057; p=0.0001). In addition, we could detect SARS-CoV-2 in three specimens in which vortexing reduced the Ct-value from 33.39 to 32.79, from 28.50 to 28.19 and from 35.58 to 32.87, respectively.

**Figure 2.**
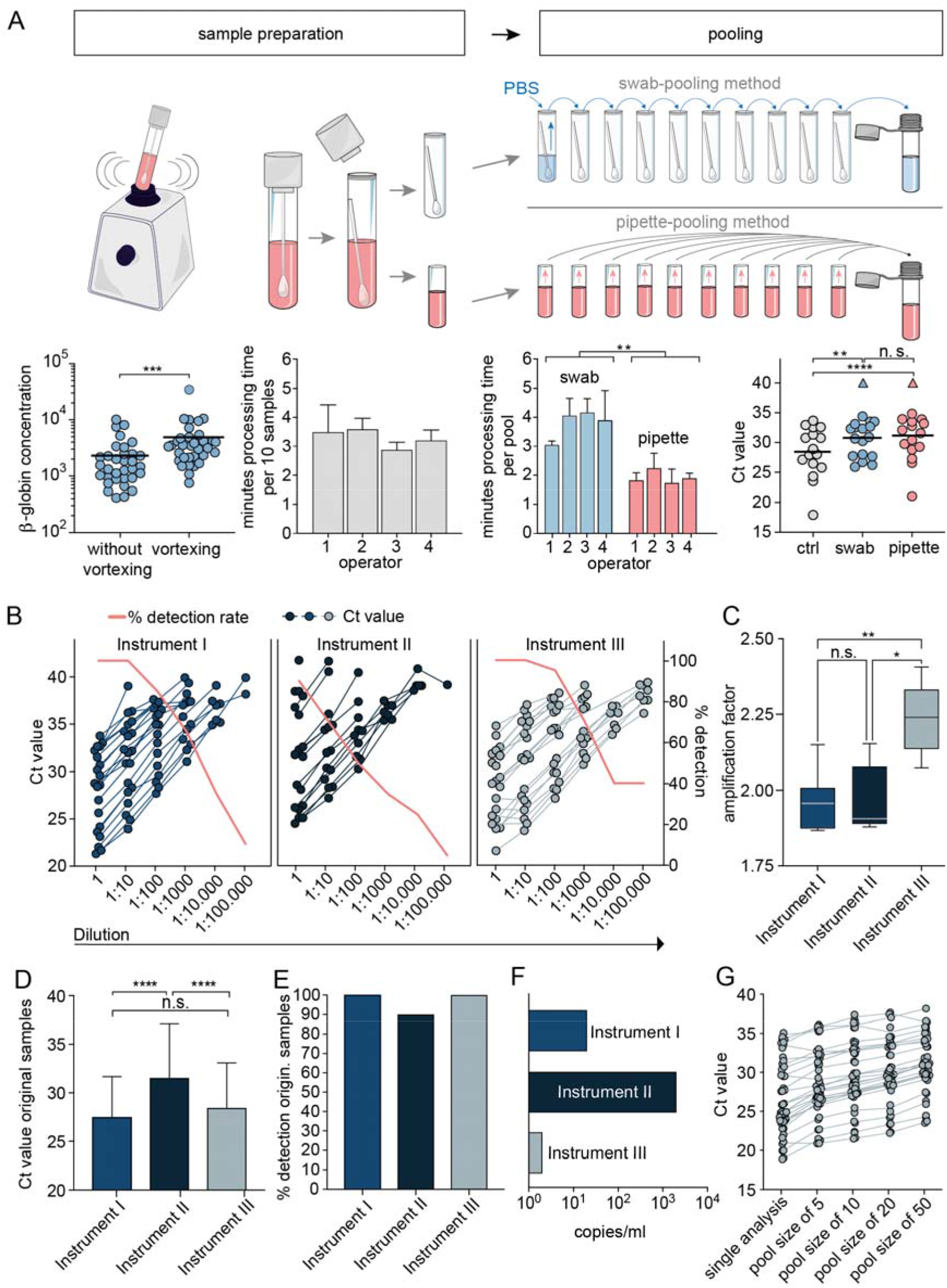
Validation of the pooling method and determining PCR sensitivity. A: β-globin concentration in individual specimens before and after vortexing (n=33). A Mann-Whitney test was performed. Sample preparation time was measured for four different operators preparing n=10 samples in 6 replicates. Swab-pooling and pipette-pooling are illustrated, and processing time was measured for four operators preparing n=6 pools with a size of 10 each (paired t-test was performed). Ct-values are displayed for n=16 single positive specimens (ctrl) as well as for each positive specimen in a pool prepared either by the pipette or swab pooling method, respectively, and tested on instrument I. Negative test results are highlighted by the triangle shape. B: Ten-fold dilution series of n=20 SARS-CoV-2-positive samples, tested with three PCR assays. C: The amplification factor was calculated for dilution series containing five Ct-values. D: The mean and standard deviation of Ct-values and E: detection rate of n=20 undiluted samples are shown. F: Lowest detectable SARS-CoV-2 copy number G: Ct-values of n=25 positive samples combined with a stock of negative specimens in a 1:5, 1:10, 1:20 and 1:50 dilution, respectively, tested on instrument III. **p ≤0.01, ***p ≤ 0.001, ****p ≤0.0001

We compared two pooling strategies. For the pipette pooling, transport medium from each of ten storage tubes were combined into one test tube. For the swab pooling, transport medium was removed, PBS added to a first tube containing the swab, vortexed, and transferred into a second swab tube followed by vortexing. After the PBS had traveled through all ten swab tubes, it was transferred into a test tube (Figure 2A). To test feasibility of both methods, four operators processed n=6 pools applying both methods, respectively. The mean processing time was 3 minutes, 47 seconds for a swab-based pool (95% CI: (2 min,59sec.)–(4min,36sec.)) and 1 minute, 55 seconds for a pipette-based pool (95% CI: (1min,33sec.)–(2min,16sec.)).

In order to investigate the sensitivity, we generated 16 different pools with each of the two pooling methods, by merging one SARS-CoV-2-positive sample with nine negative samples, respectively (Figure 2A bottom right). The mean Ct of individually tested samples was 28.41 (95% CI 26.17–30.66), 30.77 for the swab pooling method (95% CI 28.79–32.75), and 31.18 for the pipette pooling method (95% CI 28.92–33.44). There was no significant difference between Ct-values of the two pooling methods. With both methods there was a single pool, which yielded a negative test result (triangle shape in Figure 2A).

To compare the detection rates of three PCR systems used in our diagnostic laboratory, ten-fold dilution series of n=20 SARS-CoV-2-positive samples were simultaneously tested on three instruments (I, II, and III, referring to the Roche LightCycler® 480 II, the Hologic Panther Fusion®, and Roche Cobas® 6800 System). Ct-values for e-gene amplification, included in all assays, were analyzed as they yielded similar Ct-values compared to the second viral target, respectively (Supplementary Figure 1C). As shown in Figure 2B and E, instrument I and III could detect all undiluted samples whereas instrument II only detected 18 out of 20 samples. The mean Ct-values of the undiluted samples were 27.48 (95% CI 25.4– 29.57) for instrument I, 31.55 (95% CI 28.77–34.33) for instrument II and 28.44 (95% CI 26.13–30.75) for instrument III (Figure 2D). When diluting the samples 10-fold, instruments I and III could still detect all samples, whereas the detection rate of instrument II was 70%. Instrument III could still detect 95% of samples at a 1:100 dilution and showed a slower decline of the detection rate compared to instruments I and II. The amplification factors for the three instruments were 1.957 (CI 1.867-2.149), 1.906 (CI 1.879-2.153) and 2.240 (CI 2.074-2.407), respectively (Figure 2C). The lowest detectable copy number was 200 copies for instrument I, 2,000 for instrument II, and 20 copies for instrument III as determined using two INSTAND standards (Figure 2F).

To determine the detection-rate for different pool sizes, 25 positive samples with Ct-values ranging from 18.96 to 34.99 were each diluted in a stock of negative specimens and tested in duplicates on instrument III. All pools were SARS-CoV-2-positive, however, for two pools the 1:20 and 1:50 dilution resulted in one negative and one positive replicate, respectively (Figure 2G).

## 3.3 Integration of pool testing into the diagnostic routine of SARS-CoV-2 detection

The above experiments suggested the following as the optimal pool test conditions for SARS-CoV-2 detection: (a) pooling 10 samples using the two-stage hierarchical strategy; (b) vortexing the swab specimens before pooling; (c) applying the pipette-pooling method, and (d) utilizing Instrument III for PCR testing. We set up a pool testing facility, implemented features for pool testing into the laboratory software, and systematically pooled up to 488 samples per day (Figure 3A). In order to limit the number of positive samples run in pools, we preselected samples supported by algorithms of the laboratory software. Patients that had been tested positive for SARS-CoV-2 before or showed COVID-19-like symptoms were tested individually. Pool testing was performed for surveillance screenings of patients and staff of as well as for every patient admitted to the University Hospital of Cologne.

**Figure 3.**
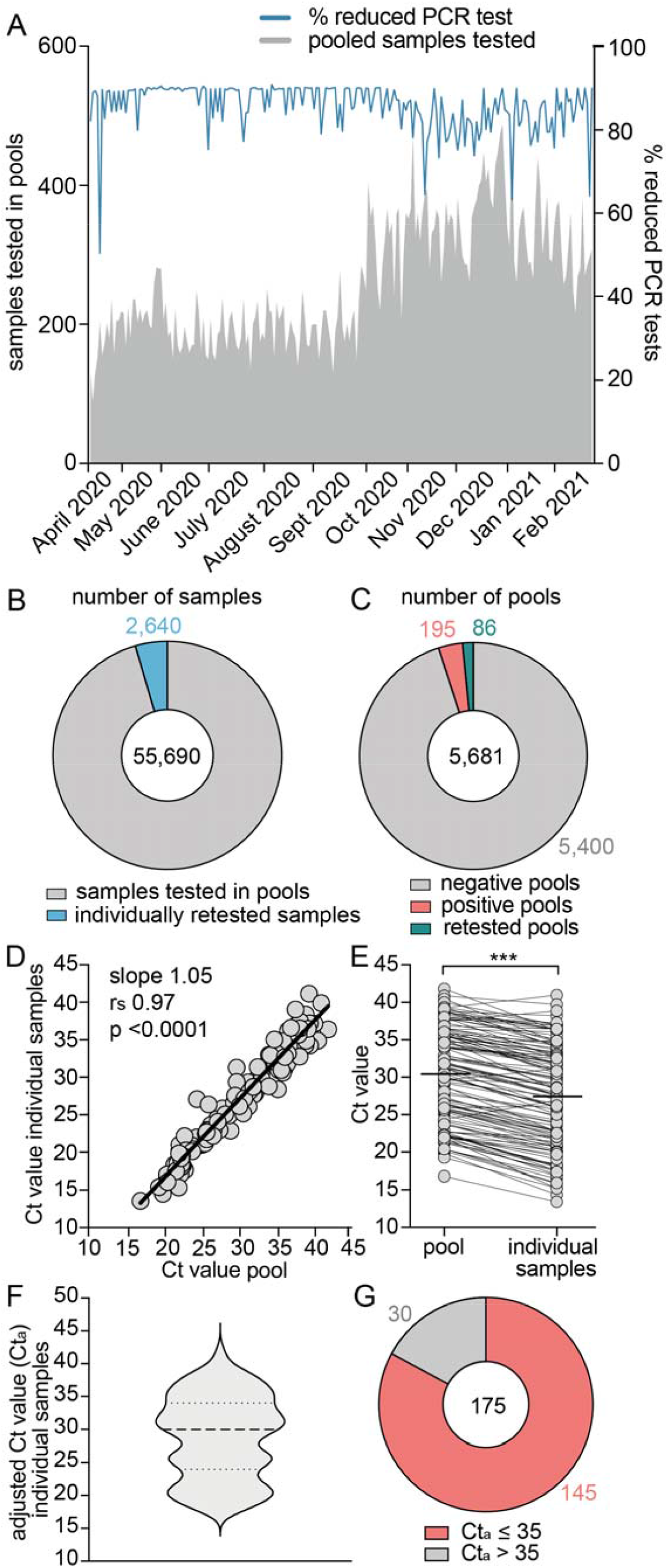
Performance of high-throughput pool testing for SARS-CoV-2 detection. A: Pool testing started on April 9, 2020. The number of pooled samples per day and the percentage of reduced PCR tests compared to individual testing (blue line) are displayed. B: The number of samples tested in pools and C: the number of pools tested during a period of 44 weeks are shown. D: Correlation of Ct-values of n=128 positive pools and the respective individual positive sample. E: Ct-values (grey dots) and mean (black line) of positive pools and the respective individual positive sample. F: Violin plot of adjusted Ct-values (Ct_a_) of n=175 individual positive samples detected in pools. Dotted lines represent quartiles and median. G: number of individual positive samples displaying adjusted Ct_a_-values ≤ 35 (red) and >35 (grey), as a Ct-value greater than 35 correlates with low infectivity (as published [12-16]).

The mean percentage of reduced PCR tests was 85.77%. Decreased savings of PCR reactions were due to retesting caused by technical issues or positive tested pools. Within 44 weeks, 55,690 samples were tested in pools and only 4.7% (n=2,640 samples) had to be retested individually (Figure 3B). As Figure 3C shows, 5,681 pools were analyzed from which 195 were positive. Another 86 pools were retested due to technical issues. In total, 47,369 PCR reactions were saved by pool testing. The Ct-values of 128 positive pools and the respective individual positive sample strongly correlated (r_s_ = 0.97, CI 0.96-0.98, p<0.0001) with a mean Ct-value of 30.39 for pools and 27.38 for individual samples (Figures 3 D and E). Ct-values of individual positive samples were adjusted to Instrument III. 82.86% of the samples displayed Ct-values ≤35 and 17.14% >35 (Figures 3 F and G).

## 4. Discussion

Large-scale testing and surveillance screenings enable the rapid detection of clusters of infections and help preventing superspreading events and uncontrolled transmission of the virus until herd immunity by vaccination is reached. However, test capacities are limited and PCR tests are cost-intensive. Pool testing is a feasible option to enable high-throughput screenings without overwhelming capacities of diagnostic laboratories. To our knowledge, this is the first systematic investigation addressing various aspects of pool testing for SARS-CoV-2 detection. However, reports on this topic have recently been published [17-31].

Eberhardt and colleagues suggest forming subgroups if a pool yields a positive result [20]. SARS-CoV-2-infected individuals need to be rapidly detected in order to apply quarantine measures and perform contact tracing. Therefore, pooling strategies need to be time efficient as well as suitable for high-throughput screenings. Following these considerations, we decided to use two-stage hierarchical pool testing with a pool size of ten samples. Increasing pool sizes, and forming subgroups after a positive result, could further improve test efficiency [20] but would delay the test result for the individual specimen. In a low prevalence setting with restricted test resources and a neglectable time aspect, for instance in developing countries, increasing pool sizes and forming subgroups is a reasonable option. Another approach is the combinatorial pool testing strategy [24,25,32]. Here, samples are assigned into multiple pools which enables the detection of infected individuals in a single round of testing. In our study, we used the hierarchical testing strategy due to logistics of a high-throughput diagnostic setting.

Pre-analytical handling can substantially impact test sensitivity, however, limited data on this topic are available. Test results are influenced by improper transport conditions, variations of the sampling device (flocked vs. cotton swabs), the transport media [33,34] as well as the anatomical structure of the pharynx (Mallampati score). We could not observe differences in Ct-values comparing oropharyngeal and combined nasal/oropharyngeal specimens. This is in line with findings of Woelfl et al., describing no differences in viral loads or detection rates when comparing nasopharyngeal and oropharyngeal specimens [35]. In addition, high viral concentrations and detection rates in salvia compared to nasopharyngeal swabs were reported [29,36] and saliva samples can be used for pool testing [37].

In our study, different operators performed specimen collection, which could be a limitation of the data, as analyzed by Basso and colleagues. Two different operators collected 70 swabs each and a high variability of test results was observed [38]. This effect should not be underestimated.

We developed a feasible pooling procedure that can readily be implemented in diagnostic routines. The preparation for pool testing contained besides extensive technical investigations, also changes in the laboratory logistics and adaptions of the laboratory software. The data communicated here will contribute to the process of finding and implementing a consensus pool testing strategy enabling larger test capacities to effectively combat the SARS-CoV-2 pandemic.

## Data Availability

The collected data are available from the corresponding author upon reasonable request.

## Funding

This research did not receive any specific grant from funding agencies in the public, commercial, or not-for-profit sectors.

## Conflict of interest

We declare no competing interests.

## Acknowledgments

We would like to thank all members of the Institute of Virology and our colleagues of the Department I of Internal Medicine and the Institute of Medical Statistics and Computational Biology, University Hospital of Cologne for supporting our work.

## Authors’ contributions

Planned and conducted experiments (MW, DA, EH, DE, JK, GHR, IF, MS, SS, MH, ASS); conceptualised the laboratory work (MW, EH, SS, FK, EK, RK); performed/helped with data analysis and interpretation (MW, HG, MA, SS, FK); wrote the manuscript draft (MW); provided clinical data, conceptualised sample collection (FD, MA, CL); made substantial revisions to the article drafts for important intellectual content (EH, HG, SS, FK); gave final approval for publication (FK); conceived and designed the overall study (FK); contributed equally to this article (FK, SS). All authors read and approved the final manuscript.

## Notes

### Competing Interest Statement

The authors have declared no competing interest.

### Funding Statement

no external funding was received for this study

### Author Declarations

The entire study was approved by the Institutional Review Board (Ethics Committee) of the Medical Faculty, University Hospital Cologne, Germany (ethical vote no. 20-1638).

