## Supplementary Figure 1 for "Safe and effective pool testing for SARS-CoV-2 detection"

**Supplementary Figure S1**

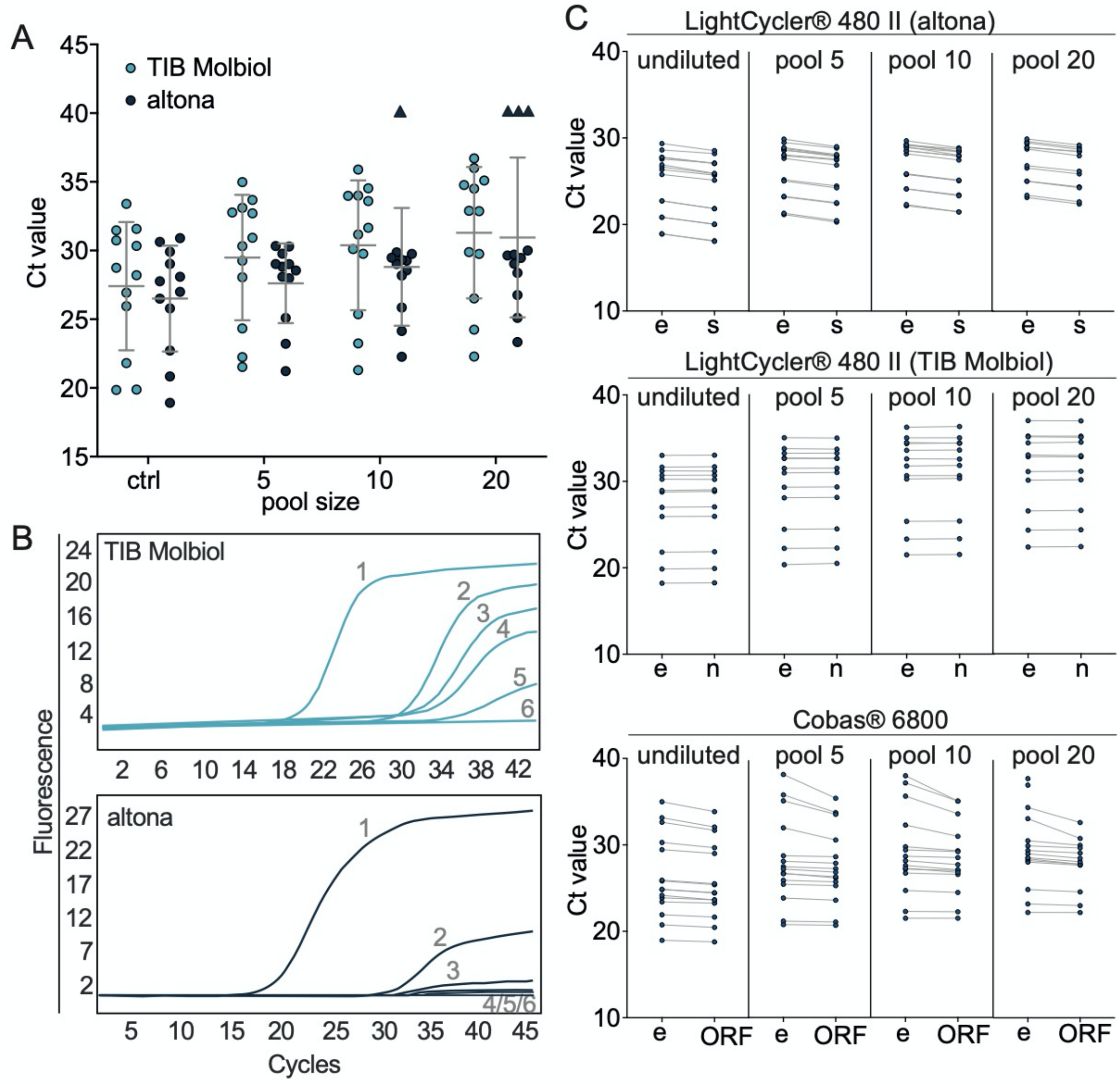


**Comparison of Ct values for different PCR kits and various viral targets.**

A: Ct values of e-gene amplification of single positive samples (ctrl) as well as pooled with negative samples of increasing sizes (duplicate reactions per point). Samples were tested using TIB Molbiol or altona kit on the LightCycler® 480 II System (n = 12). Triangles indicate negative test result.

B: Amplification curves of a positive control (1), a representative positive sample tested individually (2), as well as in pool testing with a pool size of 5 (3), 10 (4), 20 (5) and a negative control (6) using the TIB Molbiol or altona kit.

C: Ct values of positive samples undiluted and pooled with negative samples tested on the LightCycler® 480 II system (e-gene and s-gene amplification), Panther Fusion® system (e-gene and n-gene amplification) and Cobas® 6800 system (e-gene and ORF-1a/b region amplification).
