## Supplementary Figure 2 for "Safe and effective pool testing for SARS-CoV-2 detection"

Suppl Figure 2

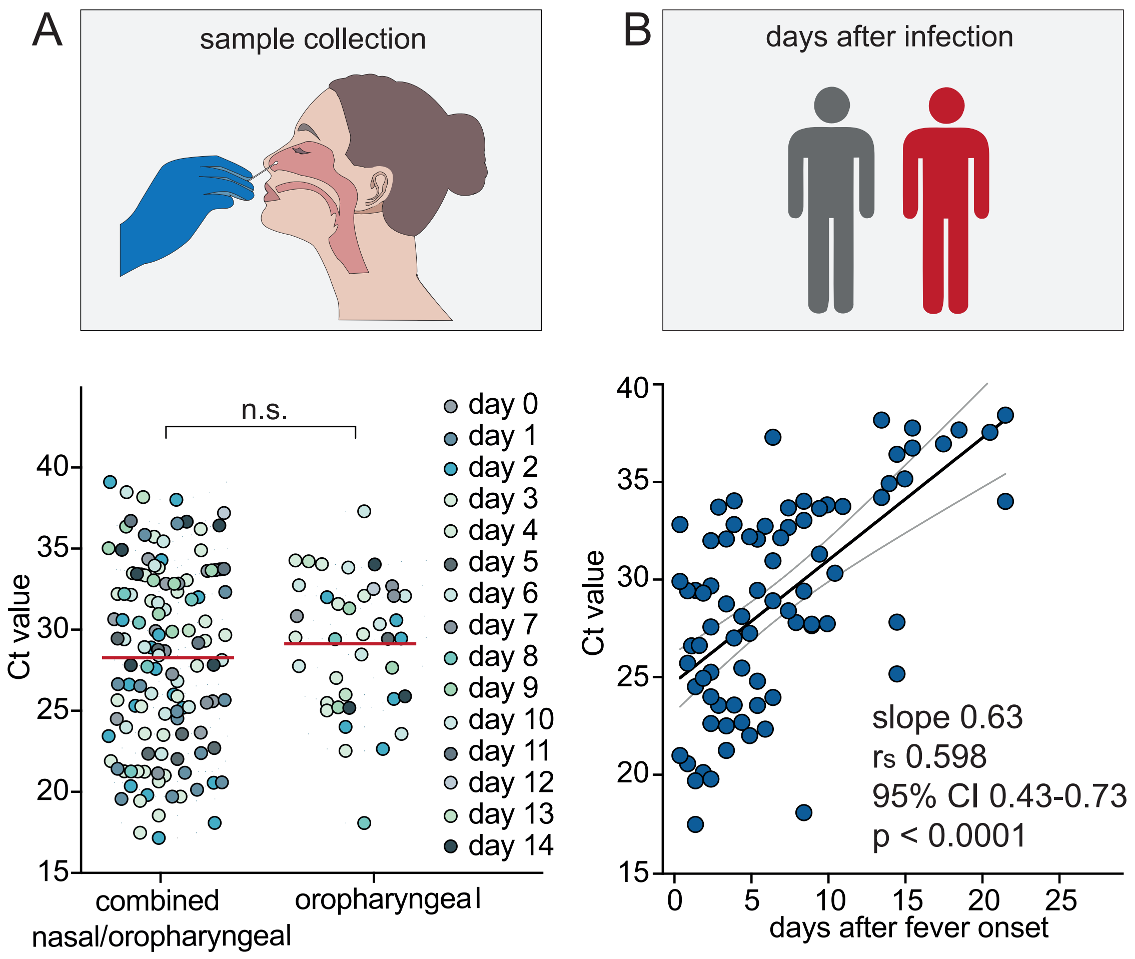


**Suppl Figure 2** Analysis of pre-analytical procedures.

A: Ct-values of oropharyngeal specimens (n=39) and combined nasal/oropharyngeal specimens (n=124). Symbols present individual specimens collected at different days after onset of initial symptoms, lines show mean. Measured on instrument III. Matched-pair analysis (paired t-test) was performed. The average time point of sample collection after onset of symptoms was 4.73 days (95% CI 4.07-5.38) for nasal/oropharyngeal swabs and 6.4 days (95% CI 5.06-7.66) for oropharyngeal swabs. Matched-pair analysis for collection timepoint after symptom onset showed similar Ct-values comparing both methods. B: Correlation between the timepoint of sample collection after fever onset and Ct-value (n=79). Measured on instrument III. The continuous line represents a nonlinear regression, dotted lines show the 95% CI.
